# Autoimmune Encephalitis, Neuromyelitis Optica Spectrum Disorder and Myelin Oligodendrocyte Glycoprotein Antibody-Associated Disease: Analysis of the Vaccine Adverse Event Reporting System (VAERS)

**DOI:** 10.1101/2024.07.15.24310444

**Authors:** Maria A. Garcia-Dominguez, Taranjit Kaur, Vincent Kipkorir, Doreen C. Cheruto, Clinton Rugut, Anosike Udochukwu, Harsimran Singh, Bahadar S. Srichawla

## Abstract

**Introduction:** Autoimmune encephalitis (AE), neuromyelitis optica spectrum disorder (NMOSD), and myelin oligodendrocyte glycoprotein antibody-associated disease (MOGAD) are complex and debilitating neurological disorders.

**Methods:** This study uses the Vaccine Adverse Event Reporting System (VAERS) to investigate the potential relationship between vaccinations and the incidence of NMOSD, AE, and MOGAD. Potential risk factors, such as age, sex, type of vaccine, and previous history of autoimmune diseases, were examined using multivariate logistic regression analysis.

**Results:** Our analysis included 161 cases: 72 NMOSD, 82 AE, and 7 MOGAD. The COVID-19 vaccine was implicated in 19/72 (26.3%) NMOSD, 43/82 (52.4%) of AE and 6/7 (85.7%) of MOGAD. The subacute temporal profile (*OR 24.4, p = 0.004*) and the presence of any comorbidity (*OR 12.49, p = 0.004*) were significantly associated with hospitalization for those with NMOSD. The subacute onset of symptoms and encephalopathy was statistically significant for hospitalization (*OR 6.15, p = 0.048*) and (*OR 10.3, p = 0.005*) respectively for patients with AE. Anti-NMDAR (N-methyl D-aspartate) antibodies were observed in 16/24 (66.7%) of AE. Treatment often involved high-dose corticosteroids or intravenous immunoglobulin (IVIG).

**Conclusions:** Most cases of vaccine induced NMOSD, AE, and MOGAD occurred secondary to the SARS-CoV-2 vaccine. The subacute onset of symptoms and the presence of encephalopathy was most associated with hospitalization.

## INTRODUCTION

Autoimmune encephalitis, neuromyelitis optica spectrum disorder (NMOSD), and myelin oligodendrocyte glycoprotein antibody-associated disease (MOGAD) are complex and debilitating neurological disorders. Each represents a distinct disease with its own pathogenesis, clinical presentation, and treatment strategies. While these conditions have been well studied in the context of their natural history and therapeutic interventions, the possible link between vaccinations and the onset or exacerbation of these disorders remains a topic of uncertainty [1, 2].

The Vaccine Adverse Event Reporting System (VAERS) is a national system established by the Centers for Disease Control and Prevention (CDC) and the Food and Drug Administration (FDA) to monitor the safety of vaccines administered in the United States. VAERS provides an essential platform for health professionals, vaccine manufacturers, and the public to report possible side effects or adverse events following immunization. Over the years, VAERS has played a critical role in identifying and understanding rare vaccine-associated adverse events, facilitating timely interventions and updates to vaccine recommendations [3]. In recent years, with the global push for vaccination against various infectious diseases, there has been increased attention to post-vaccination adverse events. Despite this backdrop, it becomes imperative to thoroughly analyze any potential association between vaccines and the onset or exacerbation of autoimmune neurological disorders such as autoimmune encephalitis, NMOSD, and MOGAD. This manuscript aims to present an in-depth analysis of the VAERS database, focusing on reports of autoimmune encephalitis, NMOSD, and MOGAD after vaccination. We will explore patterns, temporal associations, and potential risk factors to determine the nature of any association, if present. By shedding light on these potential links, we aim to contribute to a more informed understanding of vaccine safety, confirming the importance of continuous monitoring and vigilance in the realm of immunization practices.

## METHODS

### Data Source & Study Period

The primary data source for this analysis was the Vaccine Adverse Event Reporting System (VAERS). VAERS is a passive surveillance system established by the Centers for Disease Control and Prevention (CDC) and the Food and Drug Administration (FDA) to monitor the safety of vaccines administered in the United States. The system accepts reports of adverse events and vaccine errors from healthcare providers, vaccine manufacturers, and the public. The study period spanned from the inception of the VAERS database to August 23rd, 2023.

### Case Definition

Cases of autoimmune encephalitis, neuromyelitis optica spectrum disorder (NMOSD), and myelin oligodendrocyte glycoprotein-associated disease (MOGAD) were identified using standardized Medical Dictionary for Regulatory Activities (MedDRA) terms. The VAERS database was queried based on the following built-in conditions: (1) Neuromyelitis Optica Spectrum Disorder, (2) Neuromyelitis Optica, (3) Myelin Oligodendrocyte Glycoprotein Antibody Associated Disease, (4) Autoimmune Encephalitis, (5) Autoimmune Encephalopathy, (6) Encephalitis Autoimmune, and (7) Immune Mediated Encephalitis.

### Data Extraction and Cleaning

Reports containing relevant MedDRA terms were extracted from the VAERS database. Data extracted included: VAERS ID, age, sex, year reported, location/state, vaccine, time to onset of symptoms, hospitalization status, days hospitalized, symptoms, neurodiagnostic data (e.g., electroencephalogram, magnetic resonance imaging, cerebrospinal fluid studies, etc.), treatment, and results. Duplicate reports, if any, were identified using unique report numbers and were removed. Data cleaning involved the removal of incomplete entries and those with conflicting information. Data were extracted to a Microsoft Excel spreadsheet for validation, cleaning, and coding prior to data analysis.

### Statistical Analysis

Descriptive statistics were used to summarize the demographic and clinical characteristics of the identified cases. Categorical variables were presented as frequencies and percentages, while continuous variables were presented as means ± standard deviations or medians with interquartile ranges, depending on the distribution. To assess the temporal association between vaccination and the onset of symptoms, the time interval between vaccination and the onset of symptoms was extracted. Potential risk factors, such as age, sex, type of vaccine, and previous history of autoimmune diseases, were examined using multivariate logistic regression analysis. Adjusted odds ratios (aOR) with 95% confidence intervals (CI) were calculated. All statistical analyses were performed using STATA/IC *ver 15*, R-Studio *ver 1.1.447*, Python *ver 3.8* and its associated libraries including NumPy, Pandas *ver 1.3.2,* and Matplotlib *ver 3.4.3*. A p-value < 0.05 was considered statistically significant.

### Ethical Considerations

Given the deidentified nature of VAERS data, this study was exempted from institutional review board (IRB) review. However, all analyses were performed in accordance with the ethical standards of research and data privacy. No attempt was made to identify individual records within the database. The research protocol was developed and uploaded to Open Science Framework (OSF) (*Registration DOI: 10.17605/OSF.IO/8T4Y6*).

## RESULTS

A total of 161 records were obtained for coding and analysis. 72 cases of NMOSD, 82 cases of AE, and 7 cases of MOGAD were obtained throughout the VAERS database.

### Neuromyelitis Optica Spectrum Disorder (NMOSD)

A total of 72 cases of vaccine related NMOSD were obtained from VAERS. The average age was 37.17 (range: 4-72). Cases were reported between 1998 and 2022. Gender data were available in 56 patients. 41 (73.2%) were women and 15 (26.7%) were men. The COVID-19 vaccine was involved in 19/72 (26.3%) of cases. 9/19 (47.3%) Pfizer-BioNTech, 6/19 (31.6%) Moderna and 4/19 (21.0%) Janssen COVID-19 vaccines. Non-COVID-19 vaccines involved include influenza 18/72 (25.0%), human papillomavirus (HPV) vaccine 6/72 (8.3%), hepatitis B virus (HBV) vaccine 4/72, tetanus-diphtheria and pertussis (TDAP) vaccine 10/72, meningococcus vaccine 5/72, varicella vaccine 2/72, hepatitis A virus vaccine (HAV) 1/72. 38 (52.78%) individuals were treated for vaccine related NMOSD (**Figure 1**). When specified, the most common neuroimaging findings included optic neuritis and longitudinally extensive transverse myelitis (LETM), most often involving the cervical spinal cord and sometimes the thoracic. 26/72 individuals were treated with IV steroids, 19/72 plasmapheresis, 7/72 monoclonal antibodies, 6/72 intravenous immunoglobulin (IVIG), 4/72 mycophenolate, 1/72 cyclophosphamide, 1/72 azathioprine, 1/72 hydroxychloroquine. 29/72 (70.73%) of the individuals had significant clinical improvement. Three individuals died (4.17% mortality rate). Multivariate logistic regression analysis for hospitalization was completed as a result that included covariates: type of vaccine, subacute onset of symptoms as temporal profile, and having any comorbidity showing significant results of the subacute temporal profile (*OR 24.4, p = 0.004*) and having any comorbidity (*OR 12.49, p = 0.004*). The type of vaccine was not statistically significant (*OR 1.26, p = 0.25*). (**Table 1**).

**Figure 1:**
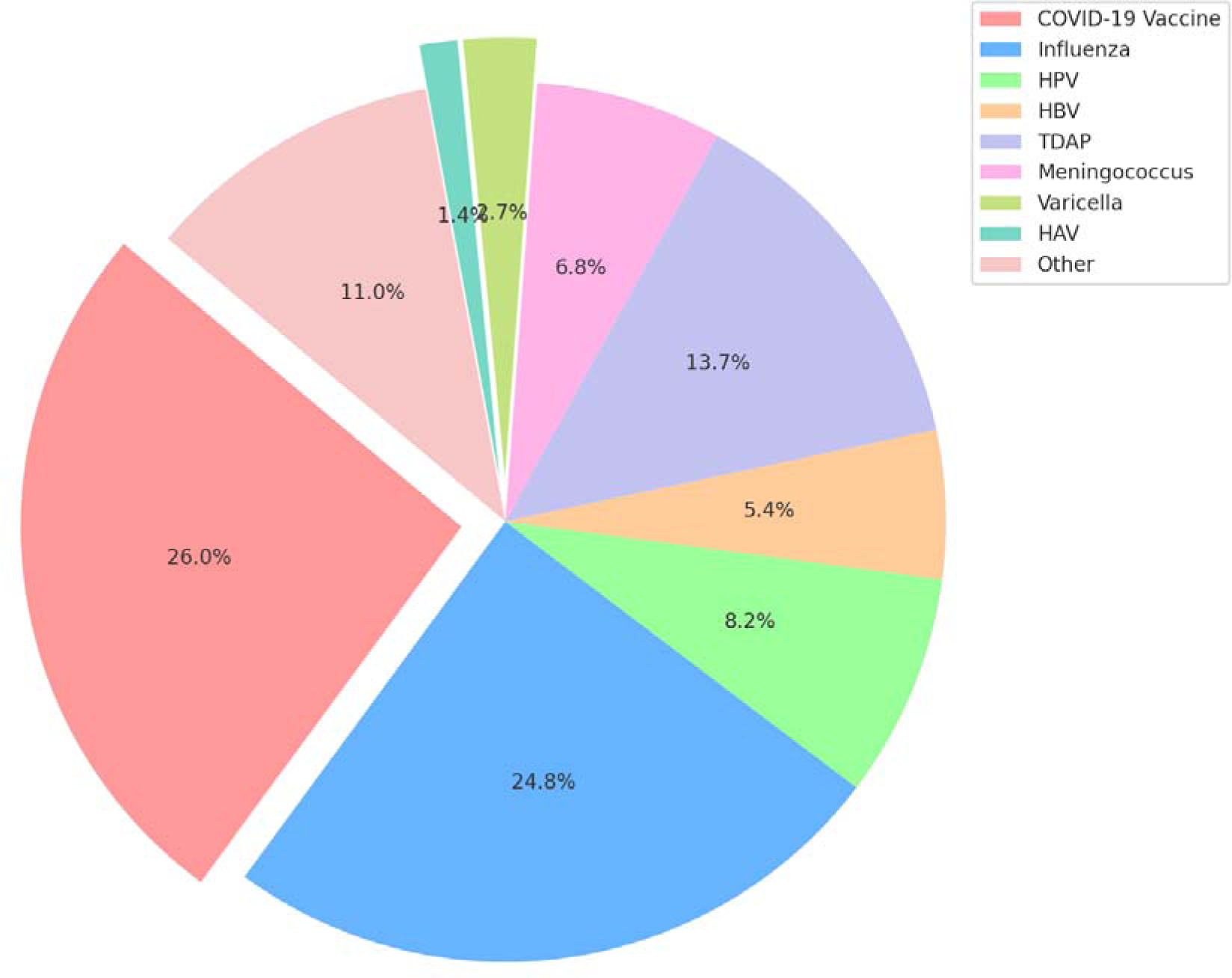
Distribution of NMOSD cases by vaccine type.

**Table 1.** Descriptive variables for patients with neuromyelitis optica spectrum disorder stratified by hospitalization status.

| Variable | Hospitalized | Non-hospitalized | p-value |
| --- | --- | --- | --- |
| **Age** |  |  |  |
| <18 | 10 | 1 | 0.41 |
| >18 | 36 | 8 |  |
| **Sex** |  |  |  |
| Male | 12 | 3 | 0.53 |
| Female | 34 | 7 |  |
| **Vaccine type** |  |  | 0.14 |
| Covid 19 | 23 | 4 |  |
| Influenza | 8 | 11 |  |
| HPV | 3 | 3 |  |
| HEP B | 4 | 1 |  |
| HEP A | 1 | 1 |  |
| TDAP | 1 | 4 |  |
| MENING B | 3 | 0 |  |
| **Comorbidities** | 21 | 3 | 0.004* |
| Neuro | 12 | 1 |  |
| HTN | 3 | 1 |  |
| Endocrinology | 6 | 2 |  |
| Allergies | 4 | 0 |  |
| **Heme/Onc** | **1** | **0** |  |
| **Rheum** | **4** | **0** |  |
| **Pulmonary** | **2** | **0** |  |
| **Recent infection** | **1** | **0** | **0.63** |
| **Temporal profile** |  |  |  |
| **Acute** | **5** | **1** | **0.38** |
| **Subacute** | **19** | **1** | **0.001\*** |
| **Chronic** | **9** | **4** | **0.53** |
| **Symptoms at presentation** |  |  |  |
| **Sensory** | **22** | **3** | **0.14** |
| **Motor** | **11** | **2** | **0.11** |
| **CN abnormalities** | **21** | **3** | **0.15** |
| **Dizziness** | **1** | **1** | **0.59** |
| **Headaches** | **5** | **0** | **0.15** |
| **Gait instability** | **9** | **1** | **0.26** |
| **Antibody present, serum** | **13** | **14** | **0.19** |
| **Treatment** | **34** | **4** | **0.87** |
| **Steroids** | **24** | **2** | **0.1** |
| **IVIG** | **6** | **0** | **0.08** |
| **PLEX** | **18** | **1** | **0.11** |
| **MAB** | **7** | **0** | **0.06** |
| **Mycophenolate mofetil** | **3** | **1** | **0.54** |
| **Improvement** | **21** | **8** | **0.24** |
| **Mortality** | **3** | **0** | **0.25** |

### Autoimmune Encephalitis (AE)

A total of 82 cases of autoimmune encephalitis (AE) were obtained from VAERS. The average reported age was 39.80 (Range: 12-89). Thirty-nine men and 41 women were identified and the gender was unknown for 2 records. The COVID-19 vaccine was involved in 43/82 (52.4%) of cases. The COVID-19 Moderna vaccine was involved 18/44 (40.1%), and the COVID-19 Pfizer vaccine 24/44 (54.5%) of cases, 1/44 was due to the Janssen vaccine, and 1 was labeled unknown. Other vaccines implicated included influenza 10/82 (12.2%), MMR 4/82, pneumococcal 2/82, anthrax 2/82 (**Figure 2**). Cases were reported between 2009 and 2023. Cases were reported most frequently in Maryland and California. 71/82 (86.9%) cases required hospitalization. A total of 24/82 (29.2%) people underwent antibody testing. Anti-NMDAR (N-methyl D-aspartate) antibodies were observed in 16/24 (66.7%). Other antibodies included anti-acetylcholine receptor, antithyroid, streptococcal, and West Nile virus antibodies. 35/82 (42.7%) people received any treatment. 9/35 individuals were treated only with high-dose intravenous methylprednisolone (IVMP), 11/35 with intravenous immunoglobulin (IVIG), 6/35 were treated with both IVIG and rituximab, one patient was treated with IVMP and rituximab. A patient was treated with IVIG and plasmapheresis. 4/35 (16%) individuals were treated with IVMP, IVIG, and plasmapheresis. The mortality rate was 4.87%.

**Figure 2:**
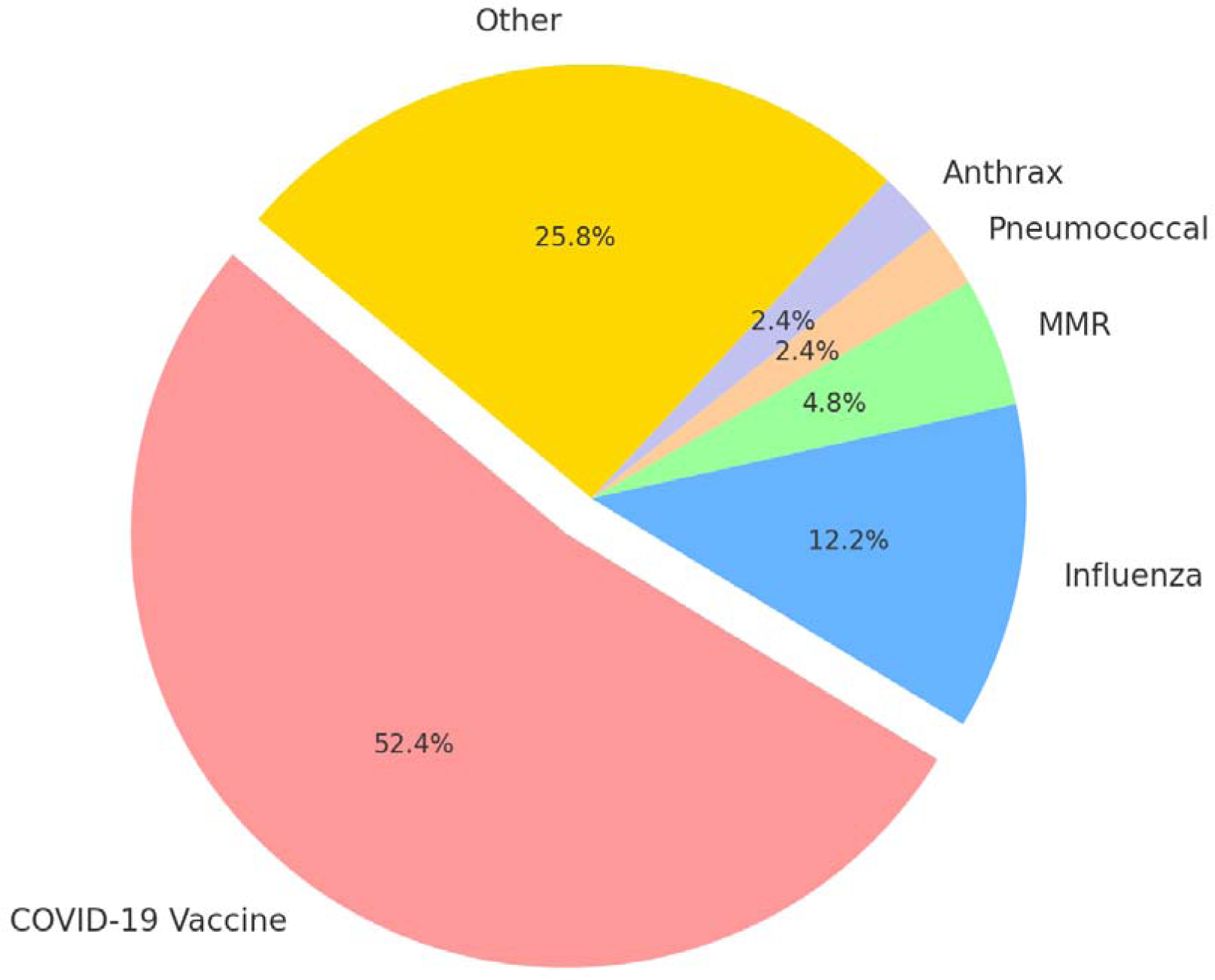
Distribution of autoimmune encephalitis cases by vaccine type.

The multivariate logistic regression analysis was completed for hospitalization as the primary outcome. Including the covariates, the temporal profile of subacute onset of symptoms and encephalopathy at presentation was statistically significant for hospitalization (*OR 6.15, p = 0.048*) and (*OR 10.3, p = 0.005*) respectively (**Table 2**).

**Table 2.**
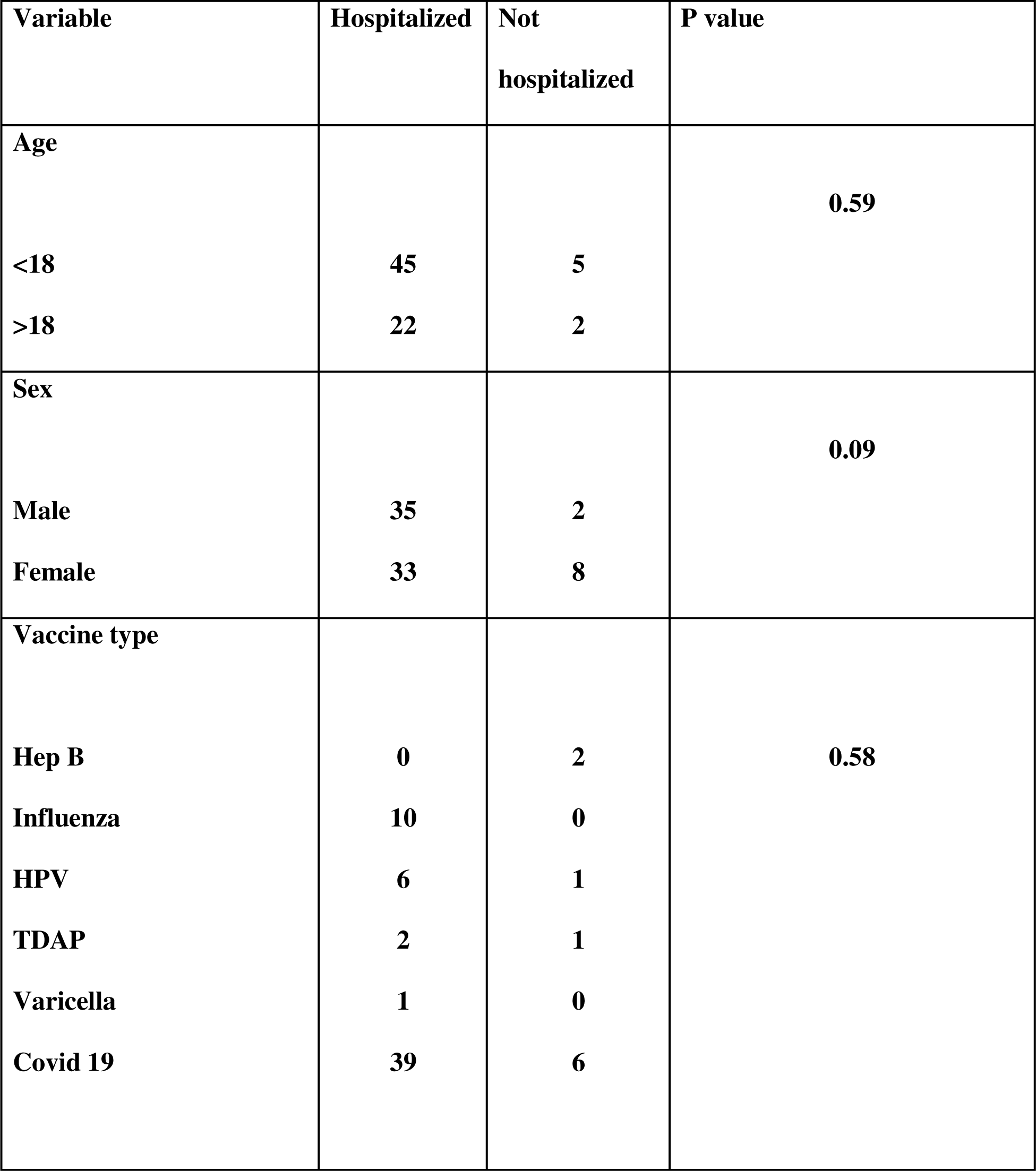
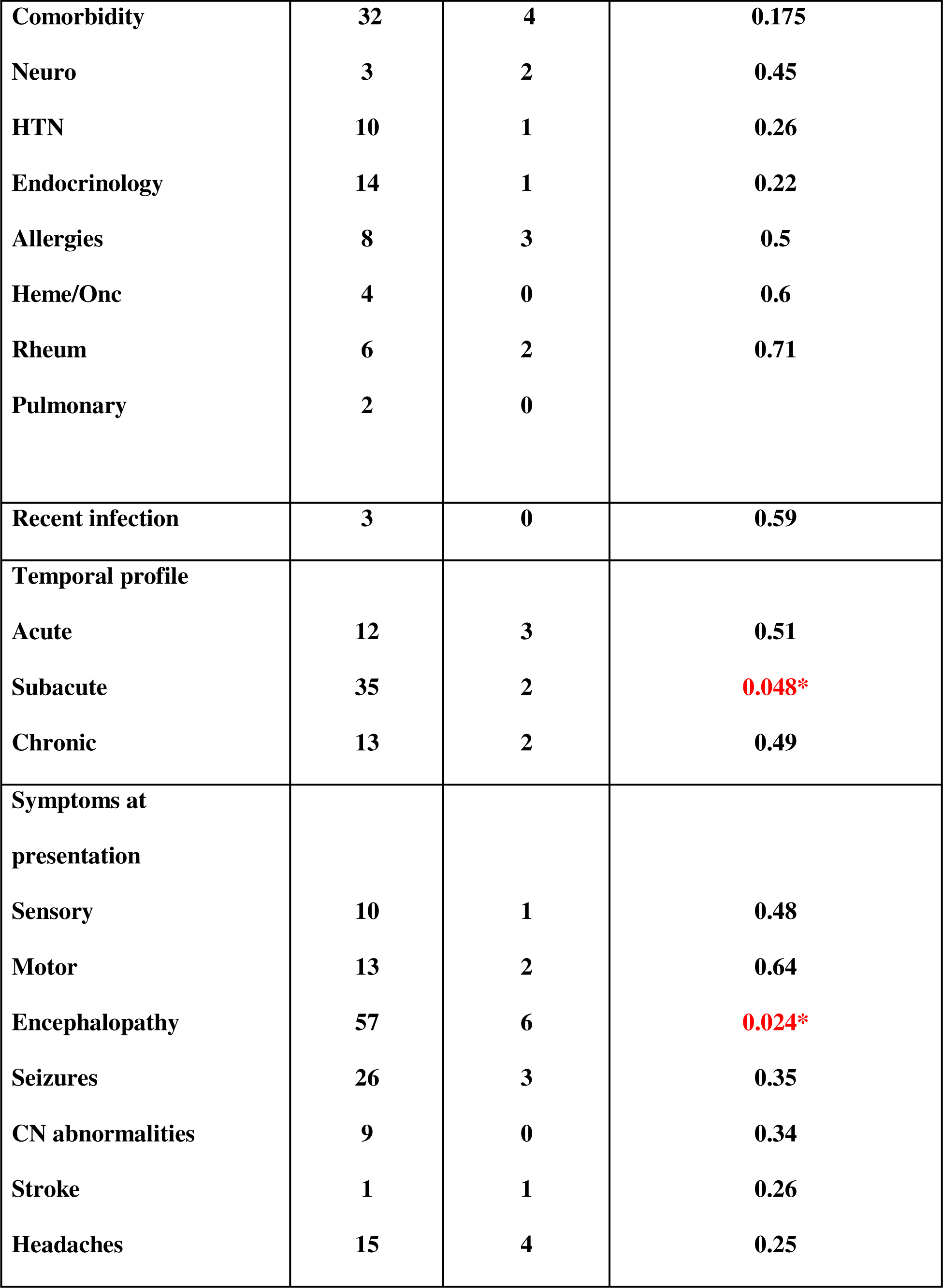
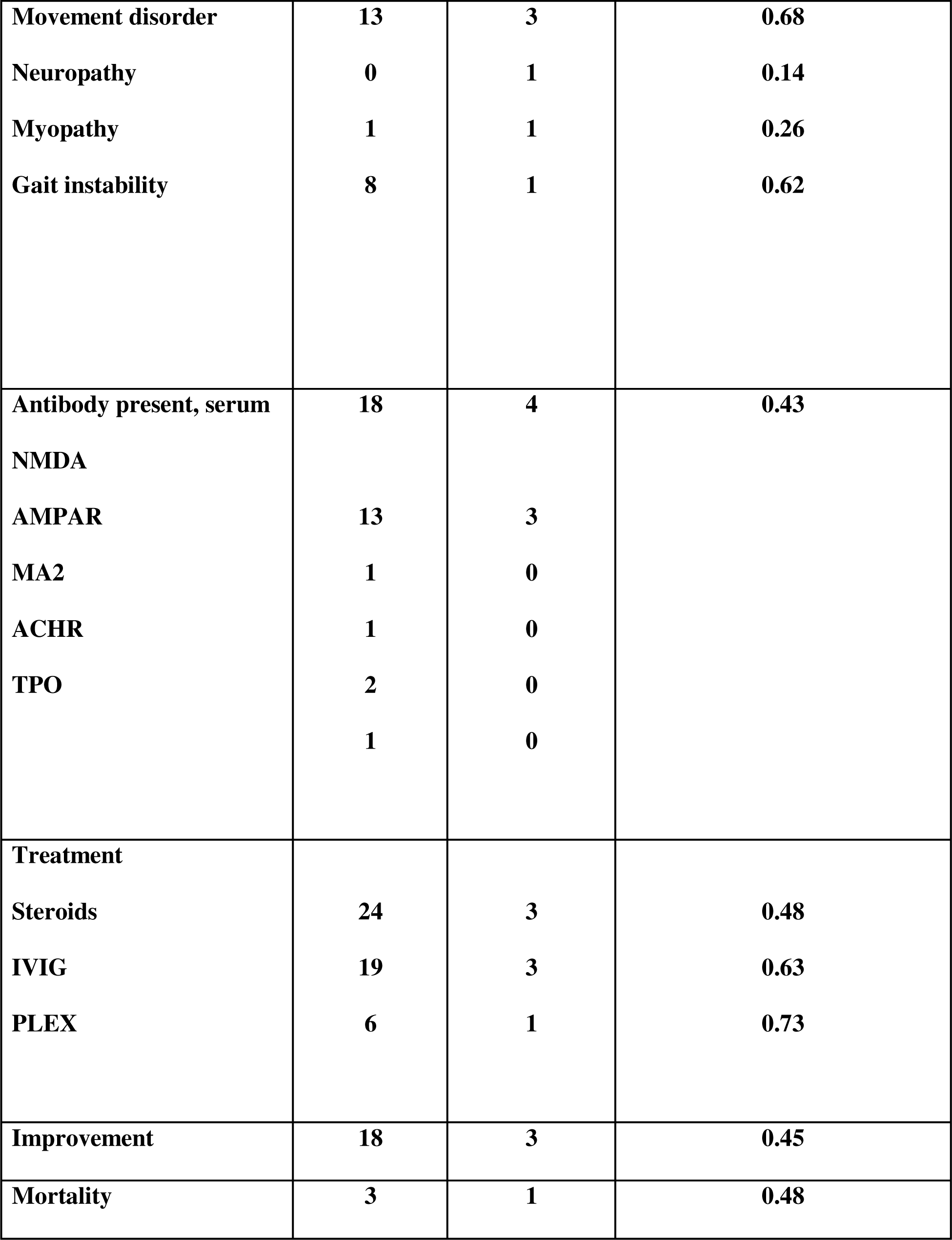
Descriptive variables for patients with autoimmune encephalitis stratified by hospitalization status.

### Myelin Oligodendrocyte Glycoprotein Associated Disease (MOGAD)

A total of 7 cases of MOGAD were obtained from VAERS. The average reported age was 45.7 (32-69). Three women and two men were identified, and the gender was unknown for 2 records. The COVID-19 vaccine was involved in 6/7 cases (3 Moderna and 3 Pfizer). There was one report of MOGAD from the varicella zoster vaccine. All cases were reported in 2021. The states involved included California, Ohio, and Texas. 5/7 required hospitalization. The median duration of hospitalization was 10 days (range: 3-23). All patients were treated with high-dose IVMP and there was no mortality.

## DISCUSSION

Autoimmune neurological disorders, including autoimmune encephalitis, NMOSD, and MOGAD, pose significant challenges in clinical management due to their complex etiologies and debilitating nature. Our study used VAERS data, a valuable resource for monitoring vaccine safety in the United States, to investigate potential links between vaccinations and autoimmune neurological disorders. Through a comprehensive analysis, our goal was to contribute to understanding vaccine safety and identification of potential risk factors associated with neurological disorders of the immune system after immunization.

In our analysis of NMOSD cases, we observed a significant proportion of individuals who reported symptoms after COVID-19 vaccination, with Pfizer-BioNTech, Moderna, and Janssen vaccines being the most involved. Other non-COVID-19 vaccines, including influenza and HPV vaccines, were also associated with NMOSD. The clinical manifestations of vaccine related NMOSD often included optic neuritis and longitudinally extensive transverse myelitis (LETM), with varying degrees of severity requiring hospitalization and diverse treatment modalities. However, only the subacute onset of symptoms was statistically significant for hospitalization in our multivariate model. Similarly, our investigation of AE cases highlighted a substantial number of reports after COVID-19 vaccination, predominantly involving Moderna and Pfizer vaccines. The clinical spectrum of vaccine-related AE encompassed a wide range of symptoms, requiring hospitalization in most cases. Similarly, the subacute temporal profile in addition to encephalopathy was associated with hospitalization. Antibody testing revealed the presence of anti-NMDAR antibodies in a significant proportion of individuals, implicating underlying autoimmune mechanisms in the pathogenesis of vaccine-associated AE. In the context of MOGAD, although the number of cases was relatively smaller, the COVID-19 vaccines were again prominently implicated, with high rates of hospitalization observed. Treatment consisted primarily of high-dose intravenous methylprednisolone (IVMP), reflecting the severity of neurological manifestations associated with vaccine-related MOGAD.

The association of a subacute temporal profile with hospitalization in cases of NMOSD and AE is noteworthy and can be attributed to the nature and progression of the symptomatology. At a subacute onset, symptoms typically develop gradually over days to weeks, potentially allowing the initial mild symptoms to intensify sufficiently to require medical intervention [4]. This progression can result in significant neurological impairment, such as optic neuritis or longitudinally extensive transverse myelitis in NMOSD, and encephalopathy in AE, which are often severe enough to require hospital-level care, including advanced diagnostics and intensive treatment modalities such as high-dose steroids, plasmapheresis, or immunotherapy [5, 6]. Consequently, patients with subacute onset are more likely to reach a threshold of severity of symptoms that requires hospitalization for acute management and close monitoring, underscoring the critical implications of temporal dynamics in these disorders.

The SARS-CoV-2 vaccine has been implicated in the triggering of various autoimmune conditions with and without central nervous system involvement [7-9]. in their systematic review, Harel et al. explore the association between NMOSD and SARS-CoV-2 infections and COVID-19 vaccinations, revealing that 41 cases emerged after exposure to the virus or after vaccination. Their analysis found a notable female preponderance and a median onset of NMOSD symptoms within 10 days after vaccination, emphasizing a possible immunological trigger related to both the infection and the vaccination processes [10]. Abdelhady et al. conducted a systematic review to explore the occurrence of encephalitis after COVID-19 vaccinations. Their review, which included 65 patients from 52 studies, found that the AstraZeneca vaccine was most associated with encephalitis, followed by Pfizer and Moderna, and most of the cases occurred after the first dose of the vaccine. In particular, the average time from vaccination to the onset of symptoms was approximately ten days, and although most patients responded well to treatments involving mainly corticosteroids and immunosuppressants, the review highlighted the lack of sufficient data to definitively confirm a causal link between COVID-19 vaccines and the development of encephalitis [11].

Vaccination can potentially trigger autoimmune responses such as NMOSD and AE due to mechanisms such as molecular mimicry and bystander activation. Molecular mimicry occurs when vaccine antigens share structural similarities with host neural antigens, leading the immune system to mistakenly attack the cells of the body after vaccination [12]. Bystander activation might also play a role, where inflammation caused by a vaccine activates dormant autoreactive T cells, which then target neural tissue [13, 14]. In some cases, adjuvants in vaccines are thought to enhance these autoimmune responses by creating a more robust immune activation, which, while generally beneficial for protection against infectious agents, may inadvertently escalate autoimmunity risks in genetically susceptible individuals [15]. Vaccinations may inadvertently influence the function of regulatory T cells (TRegs), which play a critical role in maintaining immune tolerance and preventing autoimmunity [16]. Theoretically, certain vaccine components, such as adjuvants, can induce a potent immune response that can lead to a transient dysfunction or reduction in Treg regulatory capacity [17]. This disruption might allow the activation of autoreactive T cells that would normally be suppressed, potentially initiating, or exacerbating autoimmune processes such as NMOSD and AE [18]. For example, in NMOSD, disrupted Tregs could not control autoimmunity against aquaporin 4, leading to demyelination and neurological symptoms, while in AE, insufficient Treg activity could allow unchecked inflammation within the central nervous system, resulting in encephalitic symptoms [19, 20]. These mechanisms highlight the complex interaction between vaccine components and the immune system, suggesting that while vaccines are safe and effective for the vast majority, they can, in rare circumstances, potentially initiate neurological autoimmune disorders (**Figure 3**).

**Figure 3:**
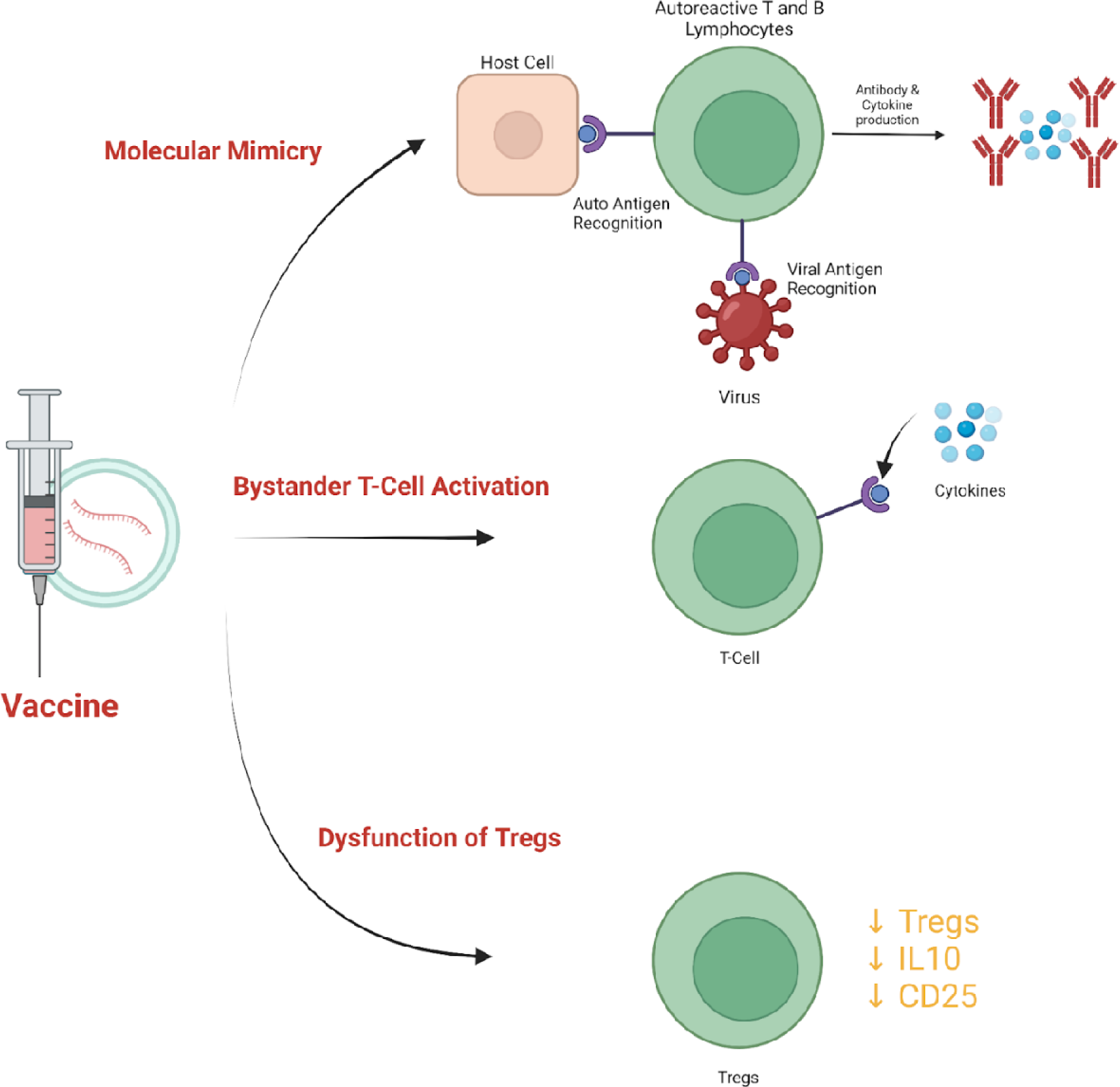
Proposed mechanisms of vaccine induced autoimmunity.

### Limitations & Future Directions

VAERS is a passive surveillance system that relies on voluntary reporting. This can lead to under-reporting or selective reporting biases, where only the most severe or the most clearly linked cases to vaccination are reported. There may be over-reporting of certain vaccines due to public concern or media attention, for example, because of the COVID-19 pandemic. Without a control group of individuals who did not receive vaccines or who did not develop adverse events, it is difficult to establish causality directly from vaccine to disease. VAERS reports can vary significantly in detail and accuracy. Some reports may be incomplete or contain errors, affecting the reliability of variables such as onset time, symptom description, and outcome.

VAERS data can establish temporal associations but are not designed to determine causality. Just because an event follows vaccination does not mean it was caused by the vaccine. The study may not adequately control for other factors that could influence results, such as underlying health conditions, concurrent medications, or demographic factors beyond gender and age. The results derived from VAERS are specific to reported cases in the United States and may not be generalizable to other populations or settings.

Future research should focus on improving vaccine safety monitoring through prospective cohort studies and improving data quality in surveillance systems such as VAERS. International collaboration could enable the pooling of data from diverse populations, increasing the robustness and generalizability of the findings. Investigating genetic markers and biomarkers that indicate susceptibility to vaccine-related adverse events could facilitate personalized vaccination strategies. Additionally, longitudinal studies are necessary to understand long-term outcomes of vaccine-induced conditions, while mechanistic studies could elucidate the underlying biological pathways, inform the development of safer vaccine formulations. Ultimately, integrating findings into healthcare policy and educational programs will be crucial to mitigate risks and increase public trust in vaccination programs.

## CONCLUSIONS

This study uses the VAERS database to investigate the possible association between vaccinations and the onset or exacerbation of autoimmune neurological disorders such as NMOSD, AE and MOGAD. Our findings suggest a notable correlation between certain vaccines, particularly COVID-19 vaccines, and the development of these conditions. However, a significant limitation to this is the positive reporting bias associated with the COVID-19 pandemic. The subacute temporal profile of the onset of symptoms emerged as a significant predictor of hospitalization, underscoring the severe impact and rapid progression of these disorders after vaccination. Our results are constrained by the limitations inherent in the VAERS system, including its reliance on voluntary reporting and its inability to establish causality. Future research should aim to enhance the robustness of vaccine safety data through improved surveillance mechanisms and prospective studies, ideally incorporating genetic and biomarker analyses to identify individuals at increased risk of vaccine-related adverse events. Understanding the mechanisms by which vaccines can trigger autoimmune responses, such as molecular mimicry and bystander activation, remains crucial.

## Statements & Declarations

### Author Contributions

**Maria A. Garcia-Dominguez:** Conceptualization, data curation, formal analysis, methodology, resources, software, validation, visualization, writing - original draft, writing - review & editing.

**Bahadar S. Srichawla**: Conceptualization, data curation, formal analysis, methodology, writing - original draft, writing - review & editing, resources, software, validation, visualization, project administration.

**Vincent Kipkorir:** writing-original draft, methodology, project administration

**Taranjit Kaur:** Data curation

**Doreen C. Cheruto:** Data curation

**Rugut Clinton:** Data curation

**Anosike Udochukwu:** Data curation

**Harsimran Singh:** Data curation

## Data Availability

All data produced in the present study are available upon reasonable request to the authors

## REFERENCES

1. Cheng MY, Ho HC, Hsu JL, Wang Y, Chen L, Lim SN, et al. Clinical Research into Central Nervous System Inflammatory Demyelinating Diseases Related to COVID-19 Vaccines. Diseases. 2024;12(3). doi: 10.3390/diseases12030060.

2. Lerusse J, Uginet M, Theaudin M, Bernard-Valnet R, Pot C, Lalive PH. [Anti-MOG associated disease]. Rev Med Suisse. 2024;20(871):828-32. doi: 10.53738/REVMED.2024.20.871.828.

3. Gee J, Shimabukuro TT, Su JR, Shay D, Ryan M, Basavaraju SV, et al. Overview of U.S. COVID-19 vaccine safety surveillance systems. Vaccine. 2024. doi: 10.1016/j.vaccine.2024.02.065.

4. Flanagan EP, Cabre P, Weinshenker BG, Sauver JS, Jacobson DJ, Majed M, et al. Epidemiology of aquaporin-4 autoimmunity and neuromyelitis optica spectrum. Ann Neurol. 2016;79(5):775–83. doi: 10.1002/ana.24617.

5. Dalmau J, Graus F. Antibody-Mediated Encephalitis. N Engl J Med. 2018;378(9):840-51. doi: 10.1056/NEJMra1708712.

6. Wingerchuk DM, Banwell B, Bennett JL, Cabre P, Carroll W, Chitnis T, et al. International consensus diagnostic criteria for neuromyelitis optica spectrum disorders. Neurology. 2015;85(2):177–89. doi: 10.1212/WNL.0000000000001729.

7. Srichawla BS. Polyarteritis Nodosa Following mRNA-1273 COVID-19 Vaccination: Case Study and Review of Immunological Mechanisms. Cureus. 2023;15(1):e33620. doi: 10.7759/cureus.33620.

8. Levi-Strauss J, Provost C, Wane N, Jacquemont T, Mele N. NMOSD typical brain lesions after COVID-19 mRNA vaccination. J Neurol. 2022;269(10):5213–5. doi: 10.1007/s00415-022-11229-1.

9. Nakano H, Yamaguchi K, Hama N, Matsumoto Y, Shinohara M, Ide H. Relapsing Anti-MOG Antibody-associated Disease following COVID-19 Vaccination: A Rare Case Report and Review of the Literature. Intern Med. 2023;62(6):923–8. doi: 10.2169/internalmedicine.0504-22.

10. Harel T, Gorman EF, Wallin MT. New onset or relapsing neuromyelitis optica temporally associated with SARS-CoV-2 infection and COVID-19 vaccination: a systematic review. Front Neurol. 2023;14:1099758. doi: 10.3389/fneur.2023.1099758.

11. Abdelhady M, Husain MA, Hawas Y, Elazb MA, Mansour LS, Mohamed M, et al. Encephalitis following COVID-19 Vaccination: A Systematic Review. Vaccines (Basel). 2023;11(3). doi: 10.3390/vaccines11030576.

12. Segal Y, Shoenfeld Y. Vaccine-induced autoimmunity: the role of molecular mimicry and immune crossreaction. Cell Mol Immunol. 2018;15(6):586–94. doi: 10.1038/cmi.2017.151.

13. Shim CH, Cho S, Shin YM, Choi JM. Emerging role of bystander T cell activation in autoimmune diseases. BMB Rep. 2022;55(2):57–64. doi: 10.5483/BMBRep.2022.55.2.183.

14. Toussirot E, Bereau M. Vaccination and Induction of Autoimmune Diseases. Inflamm Allergy Drug Targets. 2015;14(2):94–8. doi: 10.2174/1871528114666160105113046.

15. Wraith DC, Goldman M, Lambert PH. Vaccination and autoimmune disease: what is the evidence? Lancet. 2003;362(9396):1659-66. doi: 10.1016/S0140-6736(03)14802-7.

16. Rajendeeran A, Tenbrock K. Regulatory T cell function in autoimmune disease. J Transl Autoimmun. 2021;4:100130. doi: 10.1016/j.jtauto.2021.100130.

17. Keijzer C, van der Zee R, van Eden W, Broere F. Treg inducing adjuvants for therapeutic vaccination against chronic inflammatory diseases. Front Immunol. 2013;4:245. doi: 10.3389/fimmu.2013.00245.

18. Fitzgerald D, Laurent M, Funaro M, Harel A, DeAngelis T, Bangeranye C, et al. Defining the role of T lymphocytes in the immunopathogenesis of neuromyelitis optica spectrum disorder. Discov Med. 2020;29(157):91–102.

19. Byun JI, Bae JY, Moon J, Lee ST, Jung KH, Park KI, et al. Proportion of peripheral regulatory T cells in patients with autoimmune encephalitis. Encephalitis. 2021;1(3):68–72. doi: 10.47936/encephalitis.2021.00052.

20. Garcia MA, Barreras PV, Lewis A, Pinilla G, Sokoll LJ, Kickler T, et al. Cerebrospinal fluid in COVID-19 neurological complications: Neuroaxonal damage, anti-SARS-Cov2 antibodies but no evidence of cytokine storm. J Neurol Sci. 2021;427:117517. doi: 10.1016/j.jns.2021.117517.

